# Prevalence of Computer Vision Syndrome Among University Students and Faculty During COVID-19 Pandemic

**DOI:** 10.1101/2024.12.20.24319354

**Authors:** Ihab Fouad Al Ashkar, Amna Sayah Alhammadi, Ola Salam Bayram, Maria Ammari, Aisha Saleh Binashour, Ahmed Mohamed, A. Hussein, I Talaat

## Abstract

**Introduction:** The COVID-19 lockdown drastically increased screen time among students and faculty members due to online learning. Previous studies suggested increased screen time causes computer vision syndrome (CVS). This study aims to compare the prevalence of CVS among students and faculty in the UAE during the COVID-19 pandemic.

**Methods:** A descriptive cross-sectional study was conducted among 386 university students and faculty members across the UAE. A self-administered questionnaire was distributed online to collect socio-demographic data, symptoms of CVS, its associated factors, and the use of preventive measures. Informed consent was obtained, and participants who met the inclusion criteria were enrolled in the study. The indicated data were identified and analyzed using the 25th version of the SPSS program.

**Results:** Females were more likely to have computer vision syndrome, neck pain, back pain, and wrist pain. Students had a higher total screen time and a greater prevalence of Computer vision syndrome, 85.8% compared to faculty members. The prevalence of CVS was highest among students at the College of Science, 91.7%. Subjects who spent more time using the screen had more CVS symptoms than those who spent less time on screens (P= 0.039). Using laptops for more than 5 hours daily had a higher CVS prevalence of 90.6% (p=0.022). Wearing corrective eyeglasses was associated with increased CVS prevalence while taking breaks from the screen, which showed decreased prevalence. 52.5% of those with CVS experienced reduced productivity.

**Conclusion:** CVS was most prevalent in students; they also had higher screen time levels than faculty members.

## 1. Introduction

Integrating computers and smartphones into everyday life has significantly transformed how individuals interact, access information, and maintain social connections. According to recent data, approximately 5.34 billion unique mobile users, 4.70 billion active social media users, and 4.22 billion mobile social media users were reported globally [1]. These figures have continued to grow, especially spurred by the COVID-19 pandemic, which confined people indoors and drove a drastic shift from in-person interactions to digital platforms. The COVID-19 pandemic led to a dramatic increase in the adoption of digital technologies due to government-imposed lockdowns and movement restrictions. The shift enabled work, education, and socializing to continue virtually, resulting in a tenfold rise in video conferencing usage and a 30% surge in content delivery services. However, this digital transformation highlighted issues like the digital divide, technostress, and privacy concerns as societies became increasingly dependent on digital infrastructure [2].

Among the most affected groups during the COVID-19 pandemic, students and educators had to adjust rapidly to remote learning. This sudden shift to online education required students to rely heavily on digital resources such as e-books, digital lecture materials, and virtual classrooms, replacing traditional learning methods. The transition was challenging, leading to increased screen time, which research indicates contributed to various physical health issues, particularly for students. Prolonged use of digital devices has been linked to increased eye strain, neck and back pain, and other musculoskeletal disorders. Additionally, the sedentary nature of prolonged screen time has been associated with negative effects on physical health, such as weight gain and decreased physical fitness [3]. For educators, the transition to online teaching not only exacerbated existing physical health problems but also introduced new challenges, such as posture-related issues from long hours spent in front of screens [4].

Another significant health concern associated with prolonged screen time is computer vision syndrome (CVS), also known as digital eye strain, which encompasses a range of vision-related problems stemming from the extended use of computers, tablets, e-readers, and smartphones [5]. Symptoms of CVS include dry eyes, blurred vision, headaches, and neck and shoulder pain, affecting a person’s daily functioning and well-being. According to a study conducted in China, the prevalence of CVS, or dry eye disease, among high school students was reported at 70.5%, underscoring the health implications of extended screen time [6].

Given these findings, CVS has become an increasingly relevant public health issue that could affect students’ physical health, academic productivity, and quality of life. This study aims to assess the prevalence of CVS among students and faculty members in the UAE during the COVID-19 pandemic, a period characterized by an unprecedented reliance on digital devices for daily activities. Through this comparison, we seek to understand how the transition to online modes of education and work may have amplified CVS prevalence and impacted different population groups in the UAE.

## 2. Methods

### 2.1 Study design

This observational cross-sectional study measured the exposure to screen time and the prevalence of CVS symptoms in undergraduate, graduate, and postgraduate students across all majors and academic faculty in universities in the UAE between February 2021 and April 2021 during the COVID-19 pandemic. A self-administered electronic questionnaire adapted from previous literature was used for data collection [7]. A pilot study was conducted among ten undergraduate students of different majors before distribution, where the questionnaire took approximately 5 to 7 minutes to fill in. During the study period, subjects were selected using a non-probability voluntary sampling technique, and no additional benefits or payments were provided for participation. The questionnaire was created using Google Forms, in both English and Arabic, and was shared via institutional email and social media platforms.

The Research and Graduate Studies Research Ethics Committee at the University of Sharjah provided ethical approval. The study did not target vulnerable groups and was limited to adults. The gender of participants was self-reported by the participants themselves. The study ensured the privacy rights of the subjects as data was collected anonymously without identifying variables to link the information to the respondents. It was then imported to Microsoft Excel 2016 and held securely by the research team to ensure confidentiality. All participants agreed to informed consent forms, which were distributed with the questionnaire, explaining the aim of the study, and including contact details of the research group before proceeding.

### 2.2 Participants and eligibility criteria

A total of 358 self-reported male and female students and faculty members responded to the questionnaire. The subjects were 24% males, 75.9% females, 8.55% academic faculty, and 91.45% students. The estimated sample size of our population was 385 based on a 5% margin of error (ME), a 95% confidence level (CI), and an expected prevalence (P) of 50%. All university students and faculty members in the UAE who have been working and studying virtually for a minimum of 3 months were included in the study. Participants should be able to comprehend English or Arabic to respond to the questionnaire. Respondents were excluded if (1) they had a history of previous ocular surgeries or ocular pathologies, (2) they showed physical attendance to university, and (3) they were part of a hybrid attendance program resulting in a screen exposure of less than 1 hour daily.

### 2.3 Data collection

The research team developed and designed the questionnaire. It consisted of 27 questions, divided into five sections: participant demographics, CVS knowledge, CVS diagnosing criteria, participant practice, and assessment of preventive measures. For the first section, we asked about gender, age, occupation, university, and college the participants belonged to, and whether they were fully or partially enrolled in online classes. Moreover, participants were asked about their knowledge of CVS using a multiple-choice question. The criteria for diagnosing CVS were based on the presence of various symptoms and their intensity. Participants were given a chart of all CVS symptoms and had to tick the boxes if symptoms were present. The diagnosis was based on the number of symptoms the participants had. The CVS diagnosing criteria section questionnaire was retrieved from [7], p. 668.

We also asked the participants about changes in productivity, which devices they were using to attend online classes, and how long each device was used. We were also interested in knowing about the practices of the participants to see how this would impact the results. Henceforth, the participants were asked if they had consulted a physician or used eye droplets after getting the symptoms. In the last section, we were interested in knowing about the preventive measures the participants were using. This included using anti-glare screens, ergonomic chairs, taking breaks, maintaining distance, posture, etc., and using multiple choice questions to get the responses. We took the responses from the questionnaire we designed earlier for the univariate analysis. We converted them into pie charts, bar graphs, etc., to better understand the responses. For example, we used pie charts to know the gender ratio between males and females, occupation, age, etc. All the responses from the designed questions were analyzed and converted into charts and graphs using SPSS and Microsoft Excel software.

### 2.4 Data analysis

The bivariate analysis was done using the SPSS program, and it mainly focused on comparing CVS with many other important parameters we aimed to study. The prevalence of CVS was analyzed with other factors like gender, age, and occupation using a Chi-square test. Many types of tests, like the Mann-Whitney U test, Kruskal-Wallis, and others, were used to study the prevalence of CVS and the variables retrieved from the questionnaire to discover the associations.

For example, the diagnosis of CVS was analyzed, including the time spent on screens and the types of devices used (Fig. 2). Prior surgeries, eye diseases, use of protective equipment, and participant practices were also recorded and analyzed to get a clear picture of CVS and some factors that may affect its prevalence. A p-value less than 0.05 was considered significant.

## 3. Results

### 3.1 Prevalence of Computer Vision Syndrome

Among 386 participants, the prevalence of CVS between males and females was highest among females (88.7%) and lowest among males (75.3%). The chi-square test showed a p-value of 0.001, significantly lower than 0.05. Hence, the null hypothesis was rejected, and there is a significant relationship between gender and the prevalence of computer vision syndrome (Table. 1). The prevalence of CVS among different age groups showed that people aged 18 or less obtained a total count of 85 (90.4%), while people aged 19–25 got the highest percentage (84.6%). In contrast, those aged 25 and above had the lowest percentage (81.8%). The chi-square test showed a p-value of 0.284, significantly higher than 0.05. Hence, the null hypothesis cannot be rejected, and there is no difference in the prevalence of CVS among different age groups (Table. 1). Subjects with a CVS diagnosis had a longer total screen time compared to those who didn’t have CVS (Fig.1) (Mann-Whitney u value = 7658, P value = 0.039). Compared to faculty members, students had more total screen time (mean rank = 195.27, Mann-Whitney u = 5198, p = 0.303) and a higher prevalence of CVS (85.8%, p = 0.531) (Table. 1). Smartphones and laptops were the devices used most frequently (Fig. 2).

**Figure 1:**
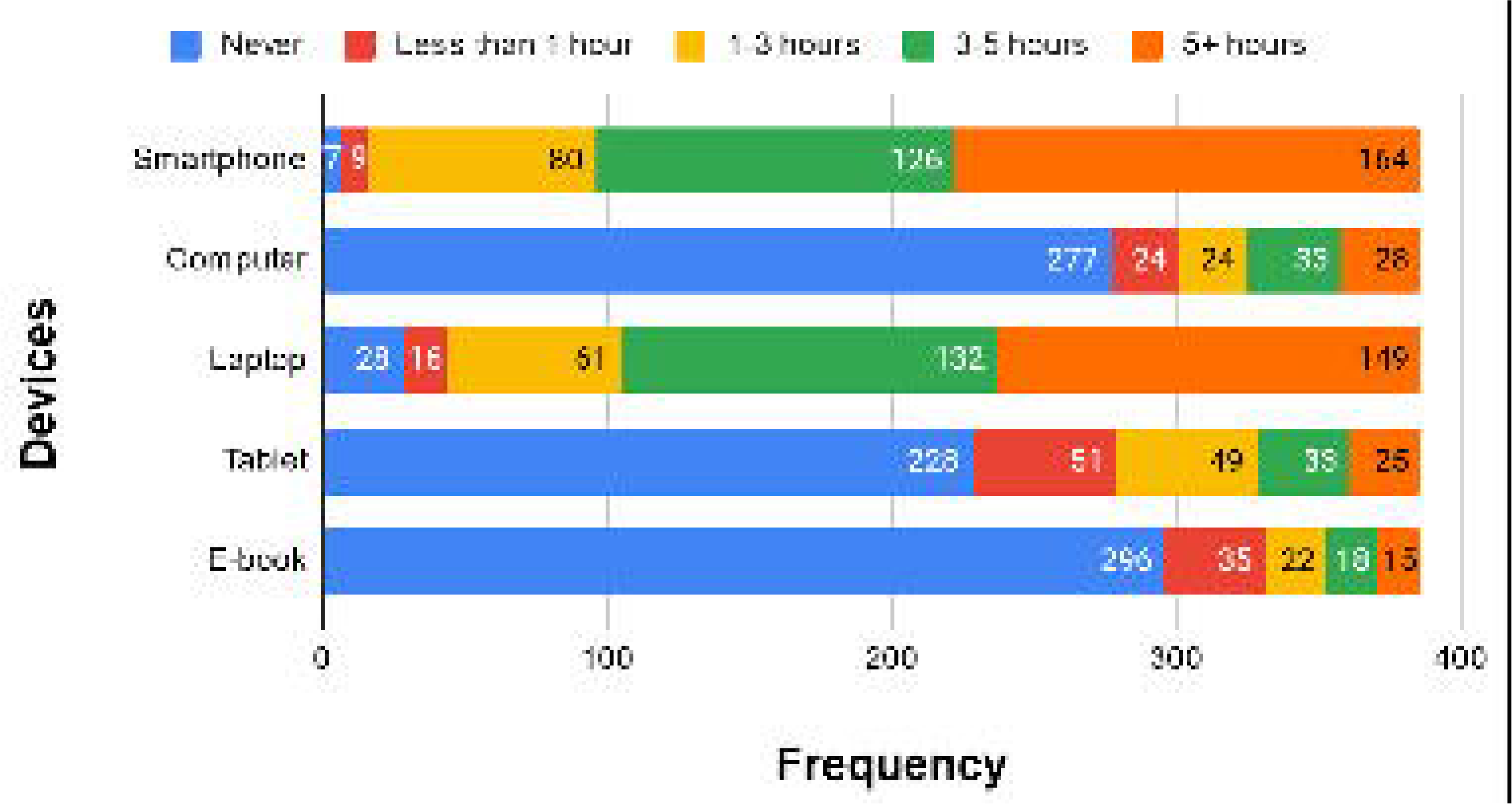
Total screen time. The figure shows the total screen time in participants with and without computer vision syndrome

**Figure 2:**
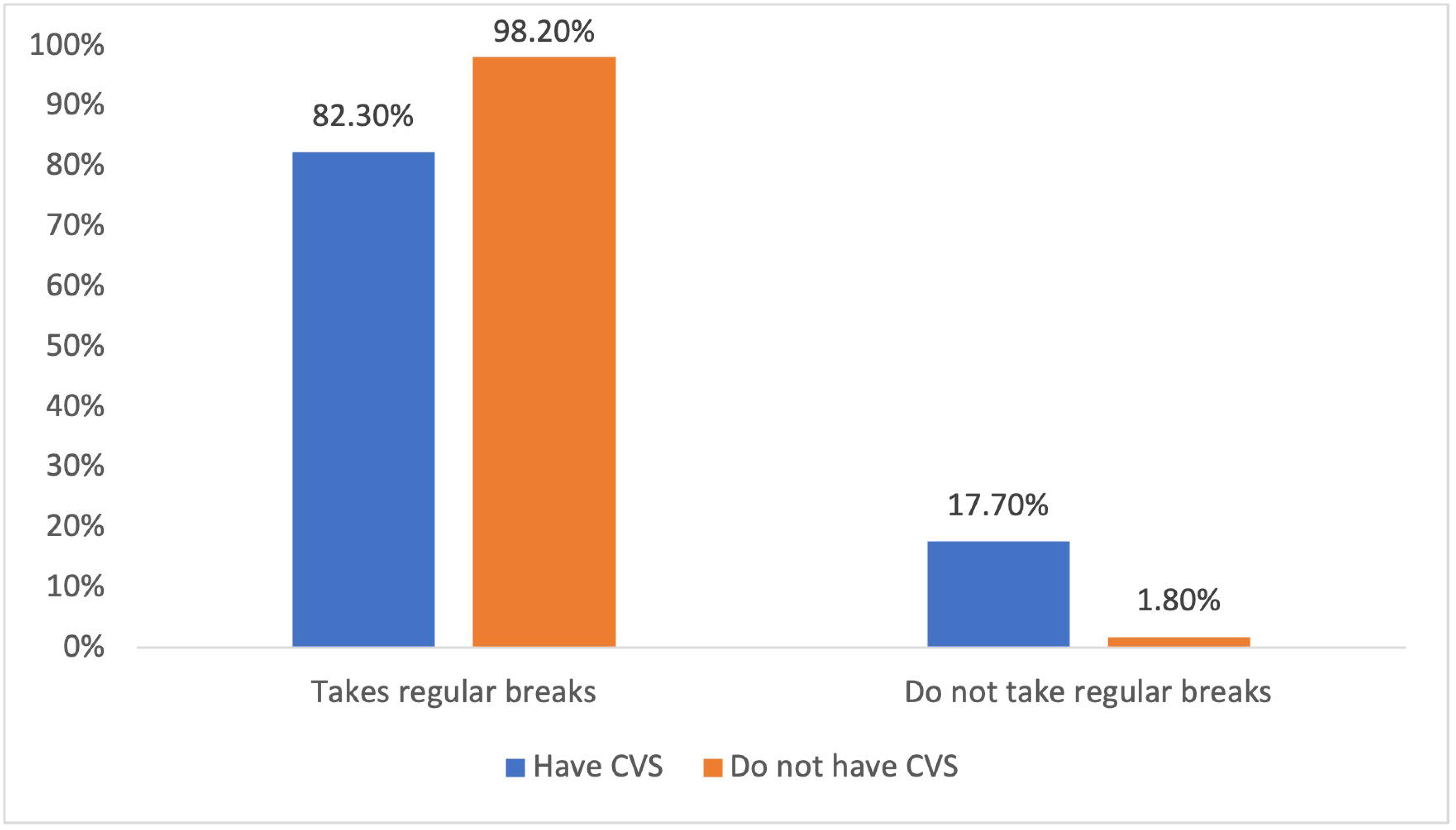
Device usage. The device usage graph shown above, presents the number of participants using different types of devices

**Table 1:** Prevalence of CVS by Demographic factors.

| Variables | N | CVS prevalence % | P-Value |
| --- | --- | --- | --- |
| Age Groups |  |  |  |
| 18 and less | 94 | 90.4% | 0.284 |
| 19 - 24 | 246 | 84.6% |  |
| 25+ | 44 | 81.8% |  |
| Sex |  |  |  |
| Female | 293 | 88.7% | 0.001 |
| Male | 93 | 75.3% |  |
| Occupation |  |  |  |
| Student | 353 | 85.8% | 0.531 |
| Faculty | 33 | 81.8% |  |
| College Major |  |  |  |
| Medicine, Dentistry or Pharmacy | 123 | 88.6% | 0.734 |
| Science | 24 | 91.7% |  |
| Health science | 47 | 80.9% |  |
| Law | 15 | 80.0% |  |
| Engineering | 92 | 85.9% |  |
| Fine arts | 28 | 82.1% |  |
| Others | 57 | 82.5% |  |
| Eyeglasses |  |  |  |
| Wears glasses | 196 | 92.3% | 0.000 |
| Doesn't wear glasses | 190 | 78.4% |  |
| Knowledge of CVS |  |  |  |
| Yes | 73 | 84.9% | 0.914 |
| No | 302 | 85.4% |  |
| Maybe | 10 | 90.0% |  |

The table summarizes the prevalence of computer vision syndrome by demographic data.

There was a positive correlation between the time spent on smartphones and the development of CVS. However, there is no statistical significance since the P value is higher than 0.05. Participants who used a laptop for less than one hour were diagnosed with CVS (3.9%). The percentages continuously increased to (13.6% for those who used their laptop for 1-3 hours, 34.8% for those who used it for 3-5 hours, and 40.9% for those who used it for more than 5 hours). This indicates an increase in CVS cases among those who spent more time on their laptop. We accepted the alternate hypothesis. There was a relationship between spending more time on a laptop and developing CVS. On investigating the prevalence of CVS regarding study major. The prevalence of CVS was highest among students in the College of Science (91.7%) and lowest among law students (80.0%) (Table. 1).

Given that the p-value is 0.734, which is >0.05 and statistically insignificant, we couldn’t reject the null hypothesis. Hence, there is no difference in study majors and the prevalence of CVS. Participants who knew about CVS and had the syndrome were 62 (18.8%). On the other hand, participants who were diagnosed with CVS and did not hear about it were 258 (78.4%).

### 3.2 The association between preventive methods and CVS

The results suggested that people who knew about CVS were likelier to take preventive measures and not get the syndrome. However, there was no statistical significance (p = 0.914) (Table. 1). We also found that those who wore glasses were more likely to have CVS at 92.3% (p=0.00, OR=3.32). 82.3% of those who took breaks experienced CVS symptoms (Fig. 3), whereas 98.2% of those who didn’t take breaks had a CVS diagnosis (p = 0.002). 52.5% of subjects with CVS experienced decreased productivity (Fig. 4); 80.3% (p = 0.001, OR = 5.849) and 81.8% (p = 0.007, OR = 2.311) experienced concomitant neck and back pain, respectively. We also investigated the prevalence of CVS and the use of lubricating eye droplets, and it showed that those who used lubricating eye droplets had a higher percentage of CVS than those who didn’t use eye droplets (Fig. 5). The chi-square test gives a p-value of 0.004, significantly lower than 0.05. Hence, the null hypothesis was rejected, and there was a significant relationship between the prevalence of CVS and the use of lubricating eye droplets. There was no relationship between surgery history and developing CVS. Henceforth, the null hypothesis is accepted. We compared the prevalence of CVS in relation to decreased productivity and no change in productivity. It showed that among the people with a high prevalence of CVS, more had decreased productivity (53.9%) compared to no change in productivity (46.1%). The chi-square test gave a p-value of 0.006, significantly lower than 0.05. Hence, the null hypothesis was rejected, and there was a significant relation between the change in productivity and the prevalence of CVS.

**Figure 3:**
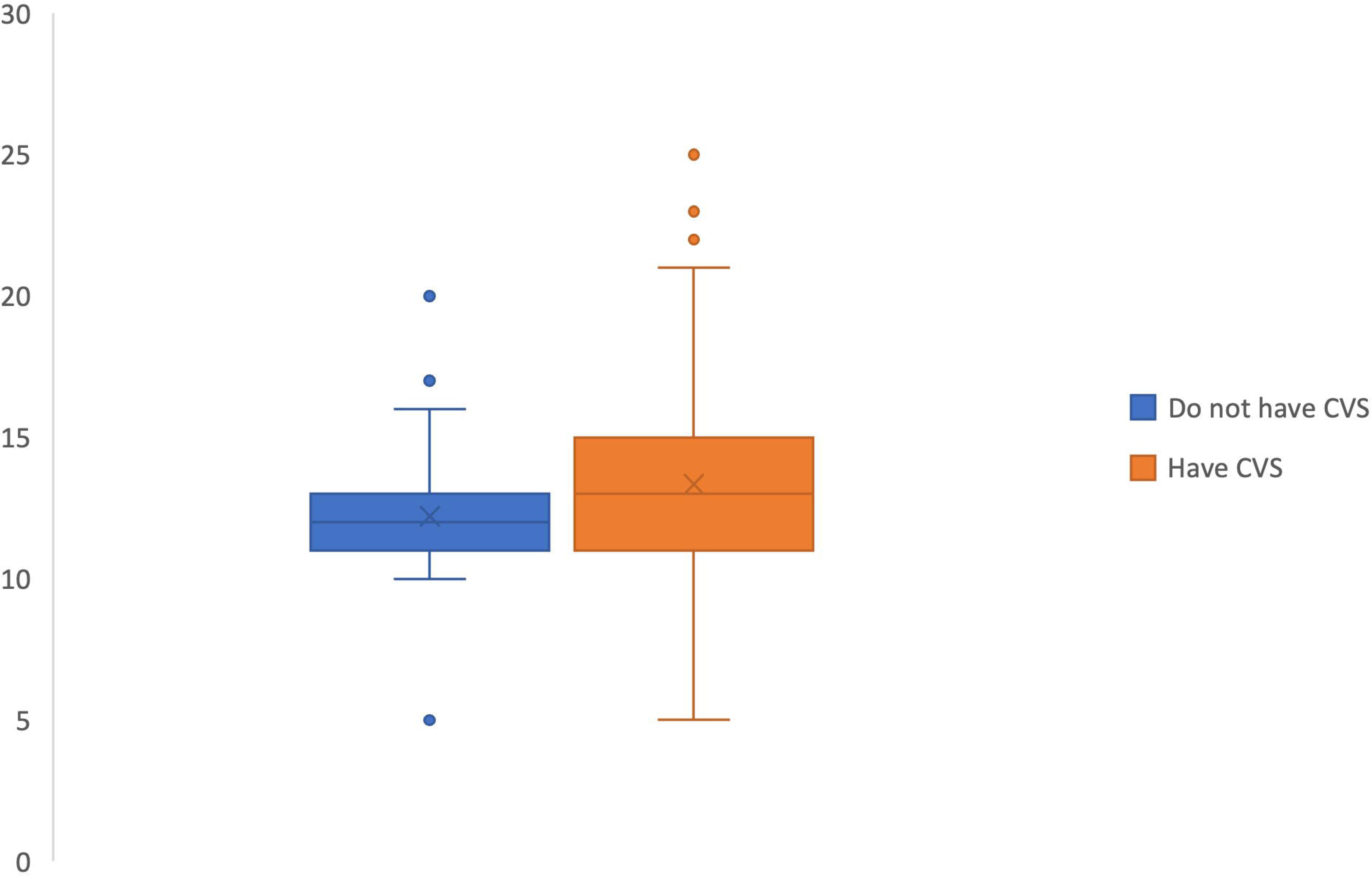
Taking regular breaks. The figure illustrates changes in productivity in participants with and without computer vision syndrome

**Figure 4:**
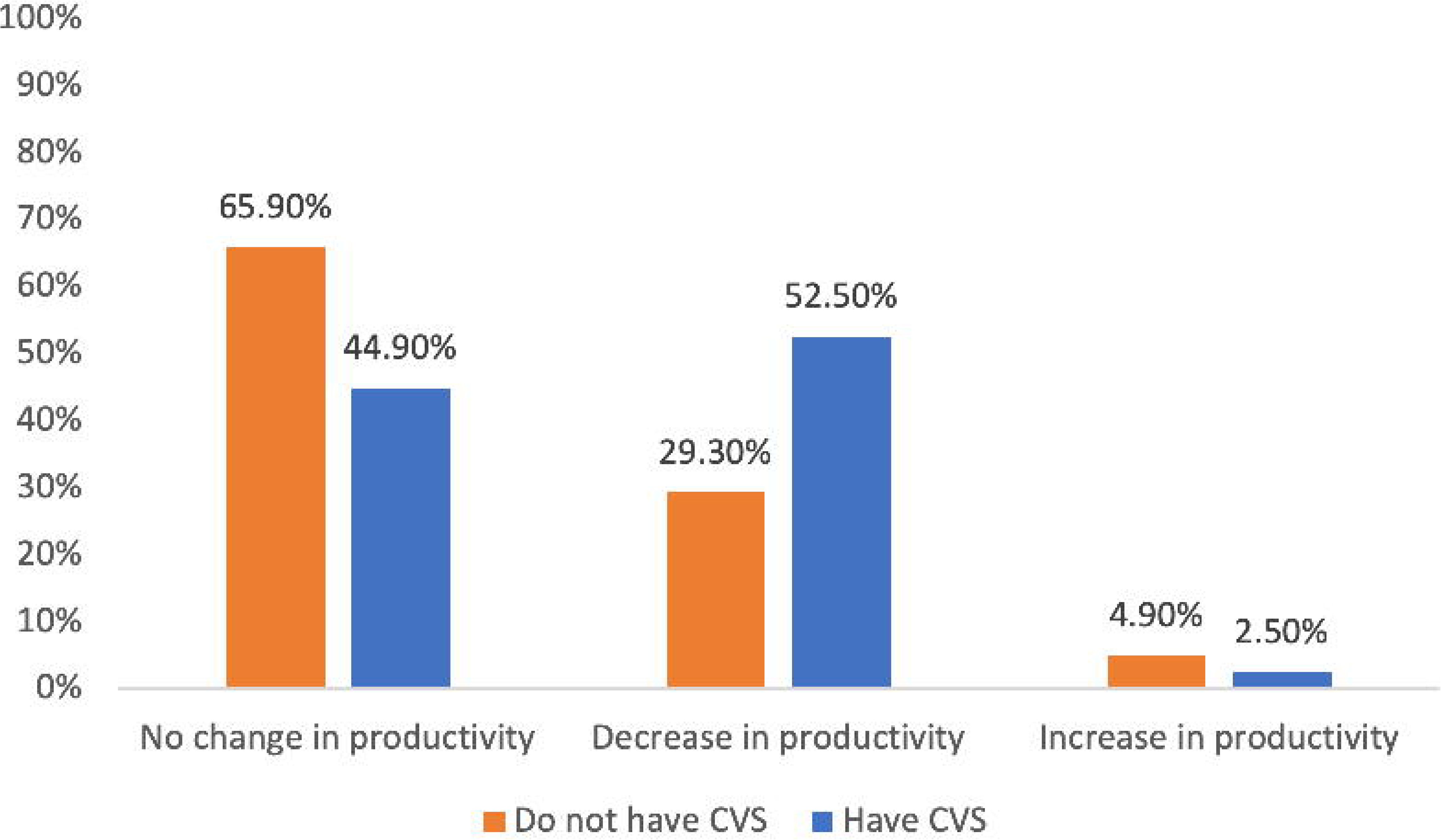
Changes in productivity. The figure illustrates changes in productivity in participants with and without computer vision syndrome

**Figure 5:**
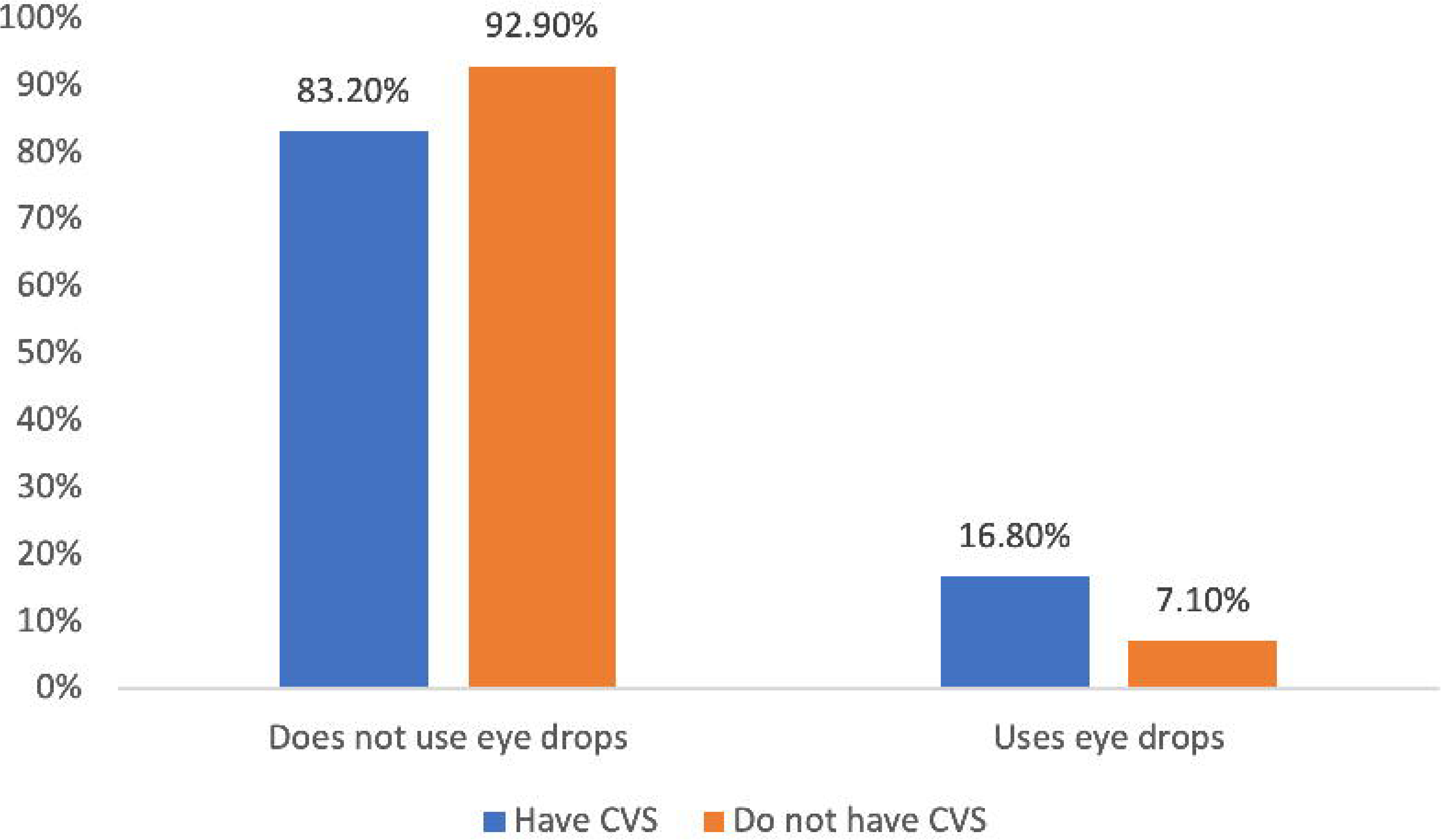
Use of eye drops. The figure shows the prevalence of computer vision syndrome in relation to the usage of lubricating eye drops

## 4. Discussion

The study demonstrated a correlation between the prevalence of CVS and increased screen exposure due to the long hours of online learning during the COVID-19 pandemic. Data was collected and studied from 386 university students and faculty members in the UAE; the data suggested that the prevalence of CVS among the participants was 85.5% during the pandemic.

The results indicated that subjects with a CVS diagnosis had a higher screen time, as the data showed a positive correlation between the time spent on laptops and the development of CVS (Fig. 1). Previous studies have proved the correlation between long periods of screen time and visual problems. A study on university students in Ajman, United Arab Emirates, reported a CVS prevalence of 72% [8]. Another cross-sectional study conducted among medical and engineering students from the suburban area of Chennai stated that a significant correlation was found between increased hours of computer use and the symptoms of redness, burning sensation, blurred vision, and dry eyes, with a CVS prevalence of 80.3% [9].

Females were more likely to get CVS along with neck, back, and wrist pain from long-term screen exposure. In addition, the participants’ knowledge of CVS in relation to its prevalence was studied. Among all the participants with CVS, only 18.8% knew about the syndrome. In comparison, the participants who did not hear about it were 78.4%, suggesting that participants who learned about CVS were more likely to take preventive measures and avoid the syndrome. Preventive techniques were also considered; participants were questioned about looking at far objects, frequent blinking, and using anti-glare screen filters, blue light filters, and ergonomic chairs as preventive techniques (Table. 2). However, no statistical significance was found between preventive techniques and CVS, unlike previous studies that stated that screens viewed at a distance had decreased the prevalence of headaches by 38%. The prevalence of tired eyes increased by 89% when screen filters were not used. The study also stated the need to increase ergonomic awareness among students and implement corrective measures to reduce the impact of computer-related vision problems [8].

**Table 2:** Patterns of device use.

| Categories |  | % (N) |
| --- | --- | --- |
| **Preventative Measures**<br>**(N=386)** |  |  |
| **Looking at far objects** | Yes | 63.20% (244) |
|  | No | 38.60% (142) |
| **Frequent blinking** | Yes | 70.40% (272) |
|  | No | 29.60% (114) |
| **Antiglare screen filters** | Yes | 88.34% (341) |
|  | No | 11.65% (45) |
| **Ergonomic chairs** | Yes | 84.97% (328) |
|  | No | 15.03% (58) |
| **Blue light filters** | Yes | 56.73% (219) |
|  | No | 43.26% (167) |
| **Screen**<br>**(N=386)** |  |  |
| **Screen Brightness** | Very Bright | 9.84% (38) |
|  | Bright | 64.51% (249) |
|  | Dull | 25.65% (99) |
| **Level of the Screen** | Above Eye level | 1.55% (6) |
|  | At Eye level | 56.22% (217) |
|  | Below Eye level | 42.23% (163) |
| **Distance from the screen** | Less than length of arm and forearm | 54.66% (211) |
|  | The length of your arm and forearm | 39.65% (153) |
|  | More than length of arm and forearm | 5.70% (22) |
| **Knowledge and breaks** |  |  |
| **Do you take Breaks? (N=372)** | Yes | 84.68% (315) |
|  | No | 15.32% (57) |
| **Frequency of breaks (N=324)** | Every 20 minutes or less | 17.28% (56) |
|  | Every 21 to 40 minutes | 30.56% (99) |
|  | Every 41 to 60 minutes | 27.47% (89) |
|  | More than 60 minutes | 24.69% (99) |
| **Knowledge of 20-20-20 method (N=386)** | Yes, and I use it | 5.96% (23) |
|  | Yes, But I don't use it | 30.05% (116) |
|  | I don't know about it | 63.99% (247) |

The table summarizes the patterns of usage of electronic devices including adherence to preventative measures. It includes most determinants and outcomes explored in the paper.

Participants who wore eyeglasses also had a high prevalence of CVS. This affected and decreased productivity. Lastly, individuals were asked if they took breaks during the long online working hours, and it was reported that those who took breaks had a lower prevalence of CVS (Fig. 3).

In line with our hypothesis, university students were more likely to get CVS than faculty members; however, as a limitation to the study, a gap between the students (91.45%) and faculty members (8.55%) sample size was reported.

## 5. Conclusion

The prevalence of CVS among students was higher compared to their faculty members. The colleges with the highest prevalence of reported CVS were the medical campuses. Most of the individuals who were diagnosed with CVS did not consult a physician. In conclusion, many of those results reflect the deceptive nature of digital transformation on students and their faculty in the education industry. Several strategies must be developed to mitigate and limit the incidence of CVS in a world of ever-changing technology.

## Funding sources

This research did not receive any specific grant from funding agencies in the public, commercial, or not-for-profit sectors.

## Data Availability

All data produced in the present study are available upon reasonable request to the authors

## Notes

### Competing Interest Statement

The authors have declared no competing interest.

### Funding Statement

This study did not receive any funding

### Author Declarations

Ethics committee of University of Sharjah College of Medicine gave ethical approval for this work.

